# Cognitive-emotional responses to ultrasonic neuromodulation of anterior cingulate cortex

**DOI:** 10.64898/2026.06.22.26356085

**Authors:** Brandon S. Cooper, Vincent Koppelmans, Thomas S. Riis, Daniel A. Feldman, Sarah Kwon, Patrick Brashear, Michael Guynn, Akiko Okifuji, Jan Kubanek, Brian J. Mickey

## Abstract

The anterior cingulate cortex (ACC) is a key brain center involved in cognitive and emotional processing that is implicated in a variety of neuropsychiatric disorders including chronic pain and depression. Circuit-targeted diagnosis and treatment of these disorders will require the capacity to precisely modulate ACC subregions. Toward that end, we recently developed and validated a novel low-intensity transcranial focused ultrasound device that can noninvasively and directly modulate ACC subdivisions in humans with millimeter precision. Here we describe the subjective reports of 36 individuals diagnosed with either chronic pain or major depression who received repeated brief stimulation trials (807 active, 797 sham; duration 30s-3min) spanning the dorsoventral extent of the ACC. Sonication immediately altered cognitive-emotional states (odds ratio 5.6, active versus sham), eliciting a positive-valence experience more often than negative (29% versus 8%) in both diagnostic groups. Sham-adjusted response rate varied across ACC targets, with the largest effects (Cohen’s d ∼ 0.8) observed in pregenual and subgenual ACC in subjects with chronic pain and depression, respectively. These rapid trial-by-trial responses to ACC stimulation predicted subsequent improvements in pain and depression severity at 24 hours. Collectively, these findings reveal that transcranial ultrasound can robustly evoke immediate, target-specific, clinically meaningful changes in cognitive-emotional state, demonstrating the potential of ultrasonic neuromodulation as a tool for individualized probing of circuit function and dysfunction.

## INTRODUCTION

The anterior cingulate cortex (ACC) serves as a central hub in a variety of physiologic neural computations at the intersection of cognition and emotion [1]. Among these functions are salience detection, autonomic control, pain affect, error monitoring, emotional regulation, and affective and reward valuation [2]. Receiving signals from distributed neural circuits representing both external and internal cues, the ACC integrates environmental and interoceptive information to regulate cognitive-emotional states [2,3]. That is, the ACC plays a critical role in shaping not only behavior but also the subjective experiential state associated with it. Accordingly, dysfunction within ACC circuitry has been implicated in a range of neuropsychiatric disorders characterized by maladaptive affective processing and impaired regulation of emotional states [4,5].

Two prevalent and persistent disorders linked to ACC dysfunction are chronic pain and treatment-resistant depression (TRD) [4–9]. In chronic pain, dorsal ACC (dACC) activity is strongly associated with the affective-motivational dimension of pain, encoding the salience and unpleasantness of nociceptive input [10]. In depression, the subgenual ACC (sgACC) is implicated in mood dysregulation and treatment resistance, with hyperactivity in this region associated with persistent negative affect [5,11–13]. Importantly, invasive neuromodulation approaches targeting these ACC nodes, including deep brain stimulation (DBS) and ablative procedures, can produce meaningful clinical improvement in both disorders, providing causal evidence that modulation of ACC activity can alter symptom-relevant affective states [14–20].

Much of what we know about ACC function in humans comes from functional neuroimaging and invasive modulation or lesioning studies [1–3]. Our ability to interrogate and modulate ACC function noninvasively has been limited. The deep midline location of ACC presents a substantial challenge for conventional noninvasive neuromodulation approaches such as transcranial magnetic or electrical stimulation, which lack the penetration depth and spatial precision required to precisely engage these circuits [21]. This limitation constrains both mechanistic investigation of ACC function and development of scalable, circuit-specific therapeutic interventions. As a result, there remains a critical need for technologies capable of noninvasively targeting ACC circuitry with sufficient focality and temporal precision to probe causal brain-behavior relationships and therapeutically modulate dysfunctional cognitive-emotional states.

Low-intensity focused ultrasound (LIFU) has emerged as a promising solution to this challenge. By delivering acoustic energy with millimeter-scale precision to deep brain structures, LIFU enables noninvasive modulation of regions inaccessible to conventional neuromodulation techniques [22–25]. We recently developed and validated a novel dual-array LIFU system which compensates for skull-induced acoustic distortion and precisely delivers deterministic ultrasound pressure to deep structures including the midline ACC [26,27]. Early human studies, including our prior investigations in chronic pain and TRD, have demonstrated that LIFU is safe and well tolerated, while producing measurable effects on neural activity, affective processing, and clinical symptoms [28–35]. These findings suggest that focal noninvasive modulation of ACC circuits may provide a powerful new approach for both mechanistic interrogation and treatment of affective neuropsychiatric disorders.

Notably, LIFU can be delivered in brief, repeated stimulation trials with high spatial and temporal precision, creating the opportunity to examine the immediate experiential consequences of focal ACC perturbation in real time. However, prior studies have largely focused on aggregate clinical outcomes measured hours to days following treatment sessions, leaving the rapid subjective effects of individual stimulation events poorly understood [36]. For invasive neuromodulation paradigms such as DBS, acute patient-reported responses to stimulation can inform targeting and parameter selection with the aim of improving longer-term therapeutic benefit [37–39]. Whether noninvasive ACC-targeted LIFU can similarly elicit immediate, perceptible changes in cognitive-emotional state remains unknown. This is a critical question because aberrant cognitive-emotional processing is a core feature of many neuropsychiatric disorders and biomarkers of LIFU treatment-response are lacking [40–42].

In the present study, we systematically characterized trial-by-trial subjective responses to brief LIFU sonications targeting ACC subregions in individuals with chronic pain and TRD. Participants underwent repeated sham and active stimulation with subsequent reporting of immediate subjective effects, enabling quantification of response valence and frequency at the level of individual sonication trials. We further examined relationships between acute response profiles and longitudinal clinical outcome measures. By focusing on rapid, within-session changes in subjective cognitive-emotional state, this work aims to provide insight into the causal role of ACC circuitry in shaping affective experience while evaluating the potential of immediate LIFU-induced responses as biomarkers of clinically meaningful target engagement and therapeutic response.

## METHODS

### Overall Study Design

Data were collected during two concurrent, pilot studies investigating the effects of LIFU neuromodulation in humans diagnosed with either chronic pain (ClinicalTrials.gov NCT05674903) [28] or treatment-resistant depression (TRD) (ClinicalTrials.gov NCT05301036) [29]. Both studies were approved by the University of Utah Institutional Review Board and all participants provided written informed consent. Previous papers described clinical outcomes and functional imaging responses [28,29]. Data reported here derive from the trial-by-trial reports of subjective experiences during offline treatment sessions, which have not been previously reported.

Both studies utilized similar double-blind, randomized, sham-controlled crossover designs involving LIFU stimulation across two designated stimulation visits (**Fig. 1**). At the first stimulation visit, participants were randomized 1:1 to receive either sham or active stimulation. This visit consisted of a 1-hour MRI session used for anatomical targeting and assessment of stimulation-related BOLD responses during concurrent LIFU and fMRI (previously reported [28,29]), followed immediately by a 1-hour treatment session outside of the MRI scanner (offline). Participants returned seven days later for a second visit in which they crossed over to the other condition and repeated offline treatment procedures. Study-specific clinical symptom measures were collected before, during and after LIFU sessions, and at follow-up visits. The chronic pain study required participants to maintain an average 24-hour pain score ≥3 (range, 0–10) to proceed to crossover, ensuring persistent symptom severity prior to the second condition. The TRD study instead used fixed crossover timing at one week. Both trials included post-stimulation monitoring through seven days. For more detailed information about the study design, please see **Figure 1** and references [28,29].

**Figure 1.**
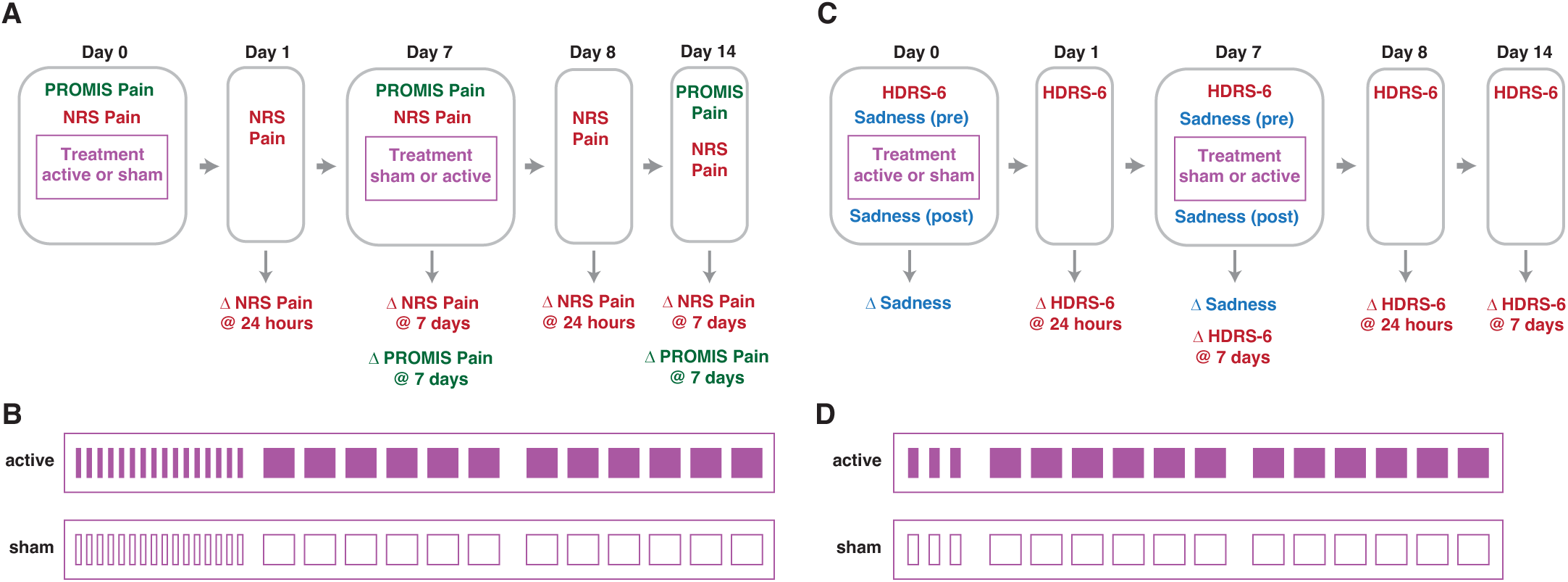
Clinical study design. **(A)** Participants with chronic pain underwent a double-blind, randomized, sham-controlled crossover protocol featuring two treatment visits in which either sham or active low-intensity transcranial focused ultrasound was delivered to the anterior cingulate cortex. Pain severity was measured with a numeric rating scale (NRS) and the Patient-Reported Outcomes Measurement Information System (PROMIS) scale. **(B)** During each treatment session, chronic pain participants received sequential trials of sham or active sonication. A block of sixteen 30-second trials preceded two blocks of six 3-minute trials. Subject reports were recorded after each trial and later scored by valence: ‘positive,’ ‘neutral,’ or ‘negative’. **(C)** Participants with treatment-resistant depression underwent a similar double-blind, randomized, sham-controlled crossover trial. Mood was measured with the Sadness subscale of the Positive and Negative Affect Schedule–Expanded (PANAS-X) and depression severity was measured with the 6-item Hamilton Depression Rating Scale (HDRS-6). **(D)** Participants with treatment-resistant depression received sham or active sonication across one block of three 60-second trials preceding two blocks of six 3-minute trials.

### Participants

Data from 19 participants with chronic pain and 17 participants with TRD were analyzed for this study. Full eligibility criteria and participant demographics are reported in the original publications [28,29]. No significant differences in demographic or clinical characteristics were found between randomization groups in either patient cohort. Four participants from the original chronic pain study and five participants from the TRD study were excluded from the current analysis because trial-by-trial subjective reports were recorded inconsistently during the early phases of the two studies.

#### Chronic pain cohort

Participants had a primary diagnosis of chronic pain present for ≥3 months. Pain etiologies were heterogeneous and included fibromyalgia (n=10), neuropathy (n=3), migraine (n=3), arthritis (n=3), and many other chronic pain syndrome co-morbidities. At baseline, the mean Brief Pain Inventory (BPI) 24-hour Numeric Rating Scale (NRS, 0–10) was 5.3 and the Patient-Reported Outcomes Measurement Information System (PROMIS) pain intensity score average was 64, corresponding to a moderate-to-severe pain severity. The sample was 60% female with mean age 46.6 years.

#### TRD cohort

Participants met diagnostic criteria for treatment-resistant depression (≥2 failed first-line therapies), currently experiencing a depressive episode without psychotic features lasting ≥2 months. Primary DSM-5 diagnoses were major depressive disorder (91%) and bipolar disorder (9%). At baseline, the mean 6-item Hamilton Depression Rating Scale (HDRS-6) score was 11.7 and the mean Quick Inventory of Depressive Symptomatology (QIDS-SR) score was 17.8, reflecting moderate-to-severe illness. The sample was 64% female with a mean age of 39.5 years.

### Randomization and Blinding

Participants were randomized 1:1 to receive sham or active stimulation at their first stimulation visit, with crossover to the alternate condition at the second visit. In each study, treatment arm allocation was concealed from participants, clinical raters, and study personnel involved in assessment and data collection. The device operator was unblinded to treatment assignment. Allocation was determined using sealed envelopes prepared in advance by an individual not involved in study procedures; envelopes were opened immediately before stimulation and revealed only to the device operator. Participants remained blinded to their randomization group throughout participation. Allocation was disclosed only after completion of the final outcome assessment conducted seven days following the second stimulation visit. Blinding integrity was confirmed in a subset of participants in the TRD cohort who were asked to guess their assigned condition [29].

### Sham Stimulation and Auditory Masking

For the sham arm of the treatment protocol, stimulation procedures were designed to mimic the auditory experience of active ultrasound without ultrasonic energy (zero pressure). Participants wore earbuds during both active and sham stimulation sessions through which an auditory mask of combined white noise and prerecorded ultrasound pulse sounds were played. These auditory stimuli were time-locked to sonication timing parameters to perceptually match acoustic cues across treatment arms.

### Ultrasound Device and Patient-specific Targeting

The neuromodulation device is described elsewhere [26,27]. In brief, low-intensity focused ultrasound was delivered using a dual phased-array transducer system mounted in a rigid frame positioned bilaterally over the temporal and parietal bones (**Fig. 2A**). Acoustic coupling gel was applied between each array and the scalp, and a thermoplastic mask immobilized the head to ensure consistent positioning across imaging and treatment sessions. Prior to stimulation, participants underwent structural T1-weighted MRI with the transducer arrays fixed in place. MRI fiducial markers enabled co-registration of the device coordinate system to individual neuroanatomy, allowing precise targeting without repositioning the participant or hardware. Treatments were then performed outside the scanner with the head secured in the identical mask-locked position to ensure reproducible targeting. The device incorporates a transmission scan and compensation algorithm to account for phase shifts, attenuation and distortion of ultrasound by obstacles on the beam path including the skull and scalp, and the dual-array design allows steering of the ultrasound focus across the ACC without shifting the subject or device frame [26,27]. The system generates an ellipsoidal focal region 20.4 × 2.4 × 3.6 mm (x, y, z) in extent. All sonications were delivered within FDA 510(k) Track 3 guidelines for diagnostic ultrasound exposure (peak intensity <190 W/cm²; spatial-peak temporal-average intensity <720 mW/cm²; mechanical index <1.9).

**Figure 2.**
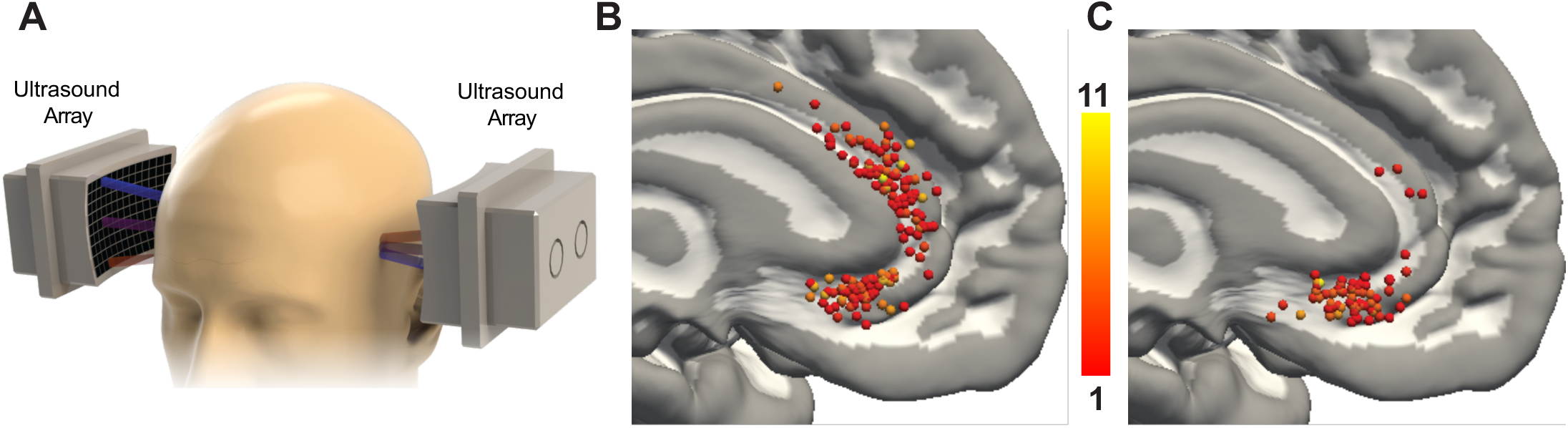
Ultrasound device and stimulation targets. **(A)** The focused ultrasound device employs bilateral phased transducer arrays over the temporal and parietal skull bones. **(B-C)** The ultrasound focus was centered on the midline and spanned bilateral anterior cingulate gyri. Sagittal images show the distribution of targets throughout the anterior cingulate cortex (inflated) mapped to MNI152 standard space in **(B)** the chronic pain cohort (518 trials) and **(C)** the treatment-resistant depression cohort (289 trials). Color represents the number of individual stimulation trials at that coordinate location.

### Stimulation Parameters

Sonication was delivered during trials lasting 30 seconds to three minutes with a 650-kHz carrier frequency and estimated peak pressure amplitude of 1.0 MPa following skull attenuation correction. Bursts were 30 ms in duration, alternating 5 ms on and 5 ms off (pulse duty cycle 50%). Intervals between bursts ranged 0.7–1.4 s (burst duty cycle 2-4%) yielding an overall duty cycle ranging 1–2%. The estimated mechanical index was 1.2, spatial peak pulse-average intensity was 31 W/cm², and spatial-peak temporal-average intensity was <720 mW/cm², meeting FDA 510(k) Track 3 guidelines for diagnostic ultrasound and consensus criteria for non-significant-risk transcranial ultrasound [31].

### Stimulation Targets

To enhance the likelihood of observing clinical effects, LIFU was delivered to multiple ACC targets in each participant. Broadly, targets were spaced approximately 4 mm apart in the sagittal plane, centered on the midline, and sonicated in random order (**Fig. 2B-C**). In the chronic pain study, eight targets spanning the subgenual (sgACC), pregenual (pgACC), and dorsal ACC (dACC) were sampled. In the TRD study, LIFU was selectively administered to 3 subregions (anterior, middle, posterior) of the sgACC. Dorsal and pregenual ACC regions were stimulated in a subset of TRD participants for exploratory purposes.

### Treatment Sessions

Treatment sessions were conducted outside the MRI scanner immediately following imaging and consisted of repeated brief sonications delivered over three sequential blocks. Individual brief stimulation trials ranged 30 seconds to three minutes and were separated by short pauses (15-60 seconds) during which participants reported immediate subjective effects related to the preceding stimulation trial. The first stimulation block (Block A) comprised trials ≤ 60 seconds. The purpose of trial-by-trial assessments during these initial 30 or 60-second sonications was to identify any stimulus locations that elicited a positive or adverse response, so that the device operator could target or avoid those locations during the later 3-minute trials. Sham and active sessions followed identical procedures, including auditory masking, except that no ultrasound was delivered with sham stimulation. Each treatment session lasted approximately 60 minutes, with 39-44 minutes total of sham or active trials (auditory mask only, or 1-2% duty cycle pulsed active LIFU) (**Fig. 1B, D**).

#### Chronic pain cohort (**Fig. 1A-B**)

Block A included sixteen 30-second sonications of 0.7-second burst intervals delivered to eight candidate targets throughout ACC in randomized order (without replacement), with each target stimulated twice. Blocks B and C each consisted of six 3-minute sonications delivered to selected targets in randomized order.

#### TRD cohort (**Fig. 1C-D**)

Block A included three to five 60-second sonications with 1.4-second burst intervals delivered across an anterior, middle and posterior sgACC. Block B included six 3-minute sonications with 1.4-second burst intervals. Block C included six 3-minute sonications with 0.7-second burst intervals.

### Collection and Scoring of Subjective Reports

Immediately following each individual sonication trial, participants were asked to report their subjective experience and perceived effect of the trial with respect to their pain/mood in an open-ended manner. Notes were taken for each sonication trial by the device operator, who was not blinded to randomization condition, with the goal of recording participant responses verbatim. This approach was deemed necessary to optimize participant safety and tolerability, given the uncertainties associated with a new brain stimulation technology. For the chronic pain cohort, numerical rating scale (NRS, range 0-10) for pain level was also solicited following individual sonication trials. Later, a rater blinded to active/sham condition scored each individual trial as either ‘negative’, ‘neutral’ or ‘positive’ in valence based on participant free-response comments and, in the case of the chronic pain cohort, changes in NRS pain score. Trials in which participants reported improvement of mood or pain were scored as positive. Conversely, trials in which mood or pain worsened were scored as negative. To be conservative with non-neutral scoring, trials during which participants referenced a tactile or audible sensation, did not mention anything overtly positive or negative about mood or pain, or simply did not comment at all were scored as neutral. For example, responses such as “felt anxious after that one,” or “annoyed,” were scored as negative, while responses such as “slightly more pleasant, on a swing set kind of happy, free,” or “much less pain,” were scored as positive. All free-response reports and valence scores can be found in the supplemental material.

### Clinical Outcome Assessments

Disorder-specific, clinically validated scales were utilized to measure changes in relevant symptoms across treatment sessions and at short-term follow-up intervals.

For the chronic pain cohort, two clinical endpoint metrics were collected. Participants completed the NRS 24-hour average pain intensity ratings on the BPI pre- and post-stimulation sessions and daily for seven days following each stimulation visit. Participants also completed PROMIS measure of pain intensity at baseline and seven days after each stimulation visit.

For the TRD cohort, two clinical outcomes were evaluated. Immediate mood change from pre- and post-stimulation session was measured by the Sadness subscale of the Positive and Negative Affect Schedule–Expanded (PANAS-X) [43] using a “right now” time frame. Depressive symptom severity was measured using the 6-item Hamilton Depression Rating Scale (HDRS-6) [44] at baseline and at 24 hours and seven days post-stimulation.

### Target Mapping

For group-level analyses, each subject’s original native-space sonication targets were translated to Montreal Neurologic Institute 152 (MNI152) common space and classified according to Yale Brain Atlas (YBA) parcels [45,46]. The YBA was selected because its centimeter-scale parcels suited our analytic goals. First, targets were normalized to the ICBM 2009a Nonlinear Symmetric template [47,48], as this template directly matches the YBA. Target normalization was accomplished by: 1) masking the native space T1-weighted MPRAGE images using Synthstrip [49] (Freesurfer 8.1.0); 2) masking the ICBM T1-weighted template image with the bundled brain mask; 3) removing bias field inhomogeneity from the skull-stripped native space image using N4ITK [50] that is bundled with Advanced Normalization Tools (ANTs) 2.5.3 [51]; 4) calculating the non-linear warp from the skull-stripped native space image to the skull-stripped template image using the antsRegistrationSyN.sh script with symmetric normalization; and, 5) applying the warp field to the native space coordinates using antsApplyTransformsToPoints. After target normalization, each coordinate in template space was then matched to a label from the YBA. Because the YBA has partial-voluming voxels that do not have a label, we used a custom growth algorithm that iteratively assigns non-labeled voxels the label of the nearest atlas label based on center of mass. Considering that targets were centered at the brain midline, sonicating both the left and right hemisphere simultaneously, coordinates appearing in both the left and right hemisphere were merged onto the left hemisphere for plotting and analysis. Group-stratified overview images showing the combined normalized ACC sonication targets of all subjects, plotted on an inflated 3D model of the ICBM template brain, can be seen in **Fig. 2B-C**.

### Analysis of Spatial Distribution of Sonication Targets

To account for participant-level differences in trial numbers of LIFU targeting tissue within a given YBA parcel, as well as individual differences in sham response rate, we calculated a sham-adjusted non-neutral response rate (ι1*r*) for each parcel for each participant. We first calculated the rate of non-neutral responses across all sham trials for each participant and used this value as the participant-specific sham response rate. We then calculated the rate of non-neutral responses to active stimulation within each parcel for each participant. From each of these values we subtracted the participant-specific sham rate, yielding a ι1*r* for each parcel and participant combination.

For each YBA region, ι1*r* scores across all participants were aggregated by averaging them. This was done to ensure that each subject contributed equally to the analysis, as the number of trials per YBA region differed among subjects (range: 1-15 (chronic pain); 1-11 (TRD)). Across both the depression and chronic pain group, this resulted in 1 to 17 observations (participants) per YBA region. For robustness, we only included regions with data for five or more subjects in our analyses. This resulted in removal of two YBA regions for the chronic pain group, and three YBA regions for the depression group. For each YBA region, one sample t-tests were used to test if the average ι1*r* scores were significantly different from zero. One-sided testing was selected because our hypothesis was that sonication elicits a response, whereas sham would not elicit a response. Alpha level thresholds were set at *p*=0.05. False-discovery-rate correction was applied to correct for multiple comparisons [52].

### Statistical Methods

Statistical analysis of participant responses was performed in R version 4.5.2 (code and specific package versions included in the supplement). Proportions of responses by valence were first analyzed with descriptive statistics. Rates of positive, neutral and negative responses by treatment type (active vs. sham) or by diagnosis (chronic pain vs. TRD) were analyzed with generalized linear mixed models (GLMM) to assess rates of non-neutral responses. Cumulative link mixed models (CLMM) were used to assess the distribution of responses by valence across conditions (order: negative, neutral, positive). To account for repeated measures and inter-individual differences in baseline responsivity, subject identity was included as a random intercept in all mixed-effects models. Cohen’s d values were calculated and are reported as standardized measures of effect size [53].

To evaluate the association between immediate response to stimulation and subsequent clinical improvement, we generated linear models with immediate positive response rate and percent change in clinical scores at the first visit, regardless of randomization order. We elected to focus on the first stimulation visit to avoid contamination in post-cross clinical data due to carryover effects seen at later timepoints. In many cases, baseline clinical scores for the second stimulation visit were too low to assess a meaningful change across or following the visit. Two participants from the chronic pain cohort and one participant from the TRD cohort were excluded from this analysis because the participants did not complete NRS or post-stimulation PANAS data, respectively.

## RESULTS

### Comparison of Sham versus Active Sonication

To evaluate the hypothesis that LIFU elicits an immediate change in cognitive-emotional experience, we compared rates of non-neutral subjective responses (positive or negative) following active ACC sonication versus sham stimulation. Across all sonication trials and all participants (chronic pain and TRD), 29% of responses were classified as positive, 63% as neutral, and 8% as negative for active stimulation (807 trials) compared to 8% positive, 88% neutral, and 4% negative for sham (797 trials) (**Fig. 3A**). A logistic regression mixed model revealed non-neutral responses were significantly more likely under the active versus sham condition (OR=5.6, 95%-CI=[4.2, 7.4], *z*=12.1, *p*<0.001, GLMM), with a bias toward positive responses (OR=2.5, 95%-CI=[2.0, 3.2], *z*=7.7, *p*<0.001, CLMM).

**Figure 3.**
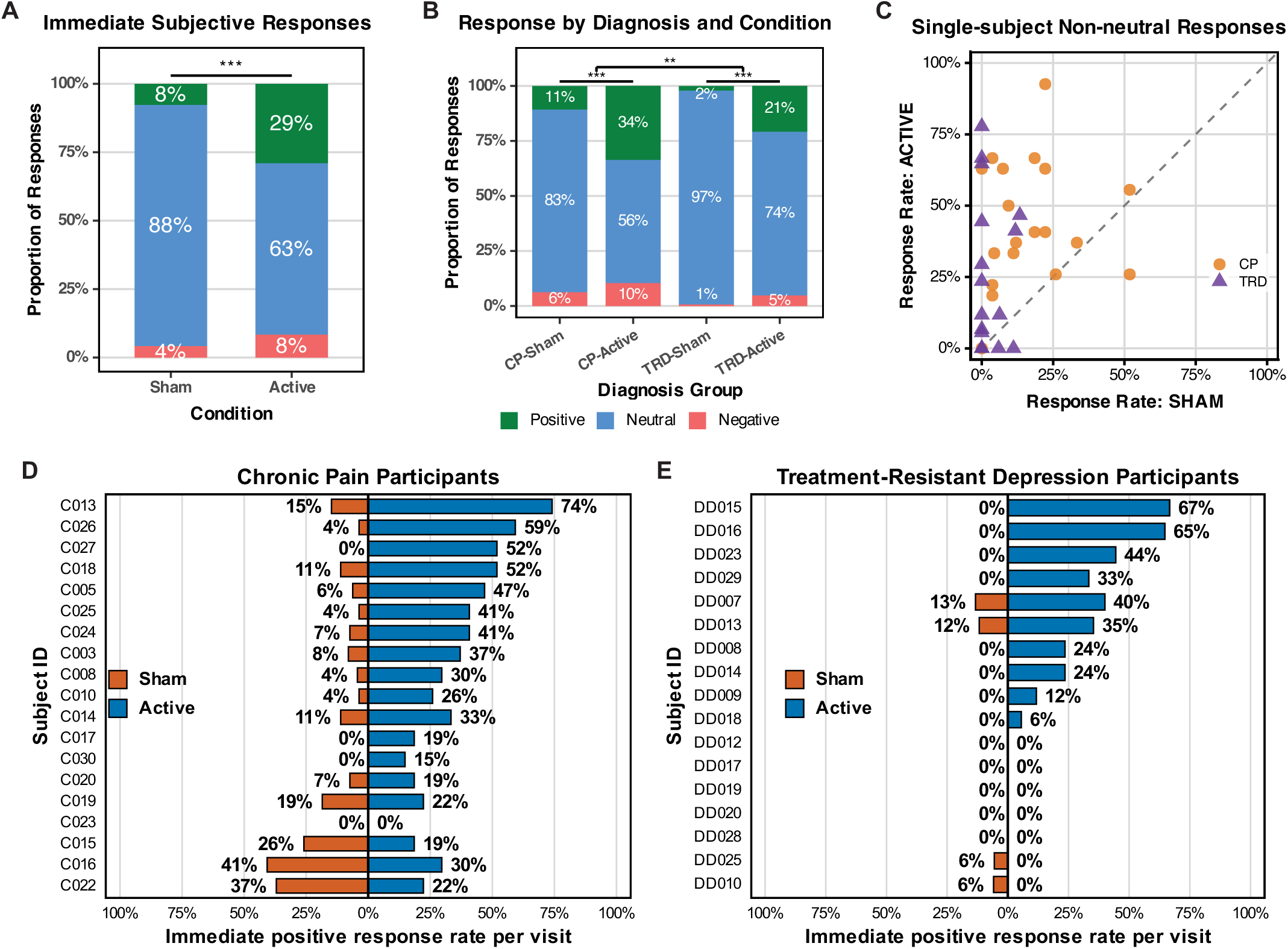
Active stimulation is more effective than sham at eliciting immediate cognitive-emotional responses. Distribution of group-level immediate subjective responses following sham or active LIFU stimulation categorized by valence **(A)** across all participants (*p*<0.001, GLMM; 797 sham trials; 807 active trials), and **(B)** stratified by diagnosis (*p*<0.001 for both chronic pain (CP) and treatment-resistant depression (TRD); *p*=0.003 for interaction of diagnosis and stimulation condition; GLMM). **C)** Scatter plot showing single subject-level non-neutral response rates of all participants (*n*=19 chronic pain, *n*=17 TRD) following sham or active LIFU stimulation. **D-E)** Butterfly plots displaying same-subject paired positive response rates to sham and active LIFU stimulation, ranked according to the difference in rate for active versus sham. **D)** chronic pain (*p*=0.001; paired Wilcoxon signed-rank test) and **E)** TRD (*p*=0.008).

We included stimulation order (active-first vs. sham-first) and its interaction with stimulation condition in the model to assess potential carryover effects. Neither the main effect of order nor the order-by-condition interaction was significant (*p*>0.35), indicating that prior exposure to sham or active stimulation did not influence subsequent immediate subjective response patterns.

As shorter-duration stimulation trials were always delivered in the first block (**Fig. 1B, D**), we also included trial duration in the model to evaluate whether response rates varied as a function of trial duration or when trials occurred within the session. Trial duration was not significantly associated with non-neutral response rates, suggesting that the observed active-versus-sham effect was not explained by block order or trial timing main effect (OR=1.26, 95%-CI=[0.96, 1.65], *p*=0.090). Similarly, in the cumulative link mixed model, trial duration was not significantly associated with the ordinal response distribution, indicating systematic shifts in response valence did not result as a function of trial timing (OR=1.06, 95%-CI=[0.83, 1.34], *p*=0.64).

### Differences by Diagnostic Group

Chronic pain and TRD participants differed in their sensitivity to LIFU. Among individuals with chronic pain, the response distribution was 34% positive, 56% neutral and 10% negative for active ACC stimulation (518 trials) compared to 11% positive, 83% neutral and 6% negative for sham stimulation (512 trials). A logistic regression mixed model revealed non-neutral responses were significantly more likely under the active versus sham condition (OR=4.4, 95%-CI=[3.2, 6.0], *z*=9.5, *p*<0.001, GLMM) and skewed towards responses with positive valence (OR=2.3, 95%-CI=[1.8, 3.1], *z*=6.1, *p*<0.001, CLMM) (**Fig. 3B**, Left).

For the TRD cohort, the response distribution was 21% positive, 74% neutral and 5% negative for active LIFU stimulation of the subgenual division of the ACC (289 trials), compared to 2% positive, 97% neutral and 1% negative for sham stimulation (285 trials). A logistic regression mixed model revealed non-neutral responses were significantly more likely under the active versus sham condition (OR=17.2, 95%-CI=[8.2, 41.2], *z*=7.1, *p*<0.001, GLMM), with a bias towards responses with positive valence (OR=3.9, 95%-CI=[2.3, 6.8], *z*=4.9, *p*<0.001, CLMM) (**Fig. 3B**, Right).

To determine whether the rate of non-neutral response to active versus sham LIFU was statistically different between diagnostic groups, we included group as a variable in the generalized linear mixed model to test for a diagnosis by stimulation condition interaction. We found a significantly greater shift in non-neutral response rate with active versus sham LIFU within the TRD cohort (*p*<0.003). The previously identified bias toward positive responses in active vs. sham was not significantly different in TRD vs. chronic pain (*p*=0.33).

Post hoc analyses revealed significantly greater sham responsivity in the chronic pain cohort than in the treatment-resistant depression cohort. During sham stimulation, chronic pain participants exhibited greater odds of reporting both non-neutral responses overall (OR=10.83, *z*=4.39, *p*<0.001, GLMM) and positive responses specifically (OR=6.19, *z*=2.94, *p*=0.003) compared with TRD participants. However, diagnosis-related differences in the overall ordered response distribution following sham-only stimulation were not significant (OR=1.20, *z*=0.74, *p*=0.46, CLMM), suggesting that the increased positive response rate may reflect greater overall responsivity rather than a selective shift toward positive responses.

### Subject-level Variation

We observed substantial variability between participants in responsiveness to LIFU. Single-subject non-neutral response rates in active and sham LIFU treatment sessions are represented for both patient cohorts in **Fig. 3C**. Response rates ranged from 0 to 92.6% under the active condition and from 0 to 51.9% under the sham condition. Several participants fall near the diagonal line of the plot (odds ratio = 1), indicating similar response rates for active versus sham condition, but the majority of participants are represented in the upper left, indicating greater responsiveness to active versus sham.

The rate of positive-valence response is plotted for each individual in **Fig. 3D-E**. The subject-level positive response rate was significantly different for active versus sham sessions for both chronic pain (*p*=0.001) and TRD (*p*=0.008; paired Wilcoxon signed-rank test). At the single-subject level, eleven of nineteen chronic pain subjects demonstrated statistically significantly elevated positive response rates to active sonication compared to sham (**Fig. 3D**, *p*<0.05, Fisher’s exact test). Four of seventeen TRD subjects demonstrated significantly elevated positive response rates to active sonication compared to sham (**Fig. 3E**, *p*<0.05, Fisher’s exact test).

In sum, these results demonstrate significant immediate effects of active LIFU sonication of the ACC on cognitive-emotional responses in participants with chronic pain and TRD, at both the group and subject levels.

### Responses Across Sonication Targets

Next, we examined how responses to active stimulation depended on LIFU target. For the chronic pain group, sham-adjusted non-neutral response rates were statistically significant in 4 regions spanning the dorsoventral range of the ACC (*p*<0.05, FDR-corrected), with the largest effect sizes (Cohen’s *d*=0.79 to 0.82) in the pregenual ACC (**Table 1** and **Fig. 4**). For the TRD group, sonication was largely restricted to three parcels comprising the subgenual ACC (**Fig. 4A-B**, parcels B-D). Sham-adjusted responses were statistically significant within the middle sgACC parcel (*d*=0.79, *p*=0.04, FDR-corrected), where the majority of sonication trials were located. (**Table 1** and **Fig. 4**).

**Table 1.** Summary statistics by Yale Brain Atlas ROI and diagnosis group. Values are averaged participant-level sham-normalized non-neutral response rates grouped by Yale Brain Atlas (YBA) ROI and diagnosis. ROI = YBA parcel label with ‘C’ indicating cingulate gyrus; Dx = diagnosis; Mean: average of the normalized participant-level responses; SD = standard deviation; p: p-value for t-test where mean>0 (one-tailed); d = Cohen’s d; n = number of participants who received at least one trial of sonication targeted to the indicated parcel FDR p = false discovery rate-corrected p-value. Bold indicates p<0.05. hidden = n<2.

| ROI | Dx | Mean | SD | p | d | n | FDR p |
| --- | --- | --- | --- | --- | --- | --- | --- |
| C<sub>B</sub> | CP | 0.15 | 0.35 | 0.094 | 0.43 | 11 | 0.138 |
| C<sub>C</sub> | CP | 0.12 | 0.38 | 0.104 | 0.32 | 17 | 0.138 |
| C<sub>D</sub> | CP | 0.25 | 0.30 | **0.014** | 0.82 | 10 | **0.040** |
| C<sub>E</sub> | CP | 0.30 | 0.37 | **0.015** | 0.81 | 10 | **0.040** |
| C<sub>F</sub> | CP | 0.28 | 0.36 | **0.010** | 0.79 | 12 | **0.040** |
| C<sub>G</sub> | CP | 0.23 | 0.38 | **0.023** | 0.59 | 14 | **0.046** |
| C<sub>H</sub> | CP | 0.16 | 0.39 | 0.121 | 0.42 | 9 | 0.138 |
| C<sub>I</sub> | CP | 0.42 | 0.65 | 0.266 | 0.64 | 2 | 0.266 |
| C<sub>J</sub> | CP | 0.05 | — | — | — | 1 | — |
| C<sub>A</sub> | TRD | 0.50 | 0.23 | 0.101 | 2.15 | 2 | 0.138 |
| C<sub>B</sub> | TRD | 0.23 | 0.24 | 0.053 | 0.93 | 5 | 0.132 |
| C<sub>C</sub> | TRD | 0.20 | 0.25 | **0.008** | 0.79 | 13 | **0.040** |
| C<sub>D</sub> | TRD | 0.34 | 0.52 | 0.110 | 0.65 | 5 | 0.138 |
| C<sub>E</sub> | TRD | -0.11 | — | — | — | 1 | — |
| C<sub>F</sub> | TRD | -0.06 | 0.08 | 0.750 | -0.71 | 2 | 0.750 |

**Figure 4.**
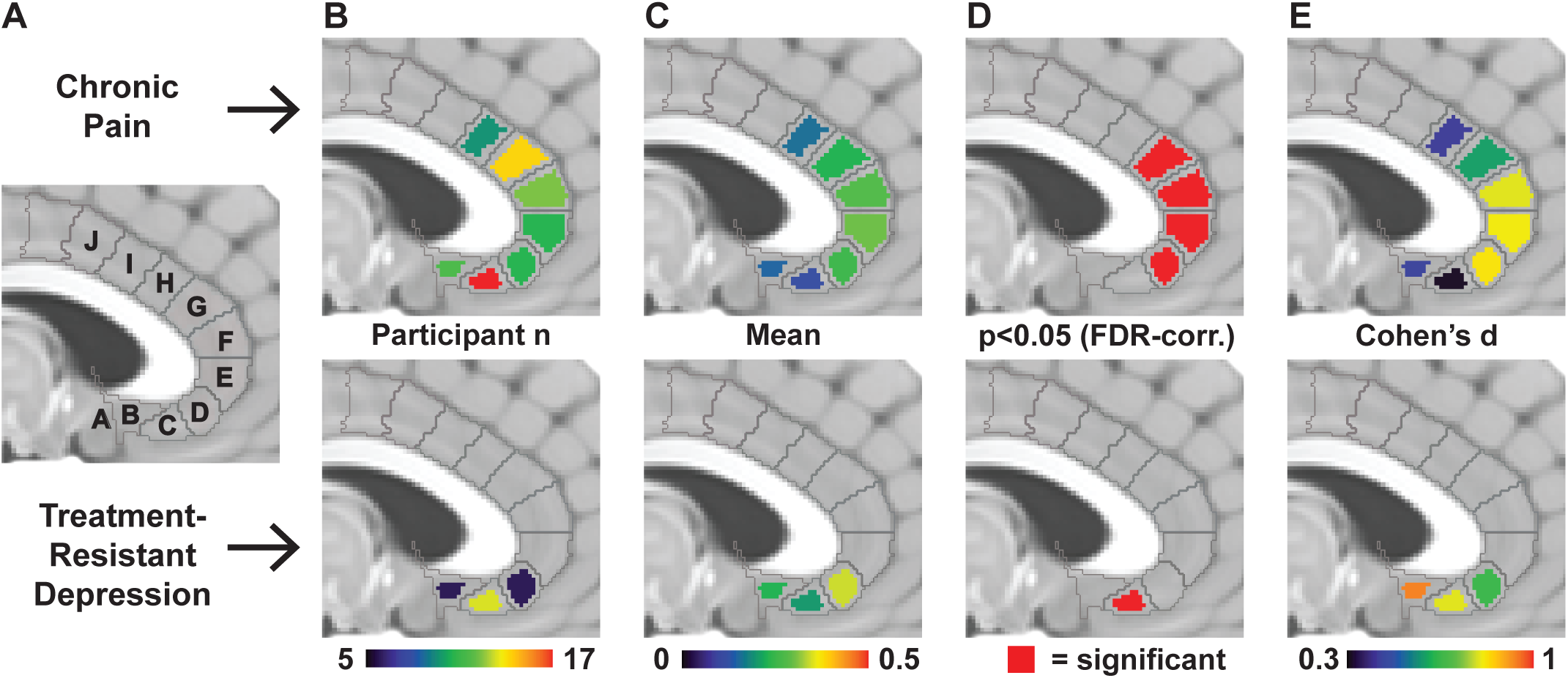
Immediate subjective responses to LIFU depend on target location. All images display standardized parcelation of the cingulate cortex on the MNI152 brain. *Top*: data from the chronic pain cohort. *Bottom*: data from the treatment-resistant depression cohort. **(A)** Labels utilized in our analyses corresponding to Yale Brain Atlas parcels (CX) within cingulate cortex (CA – CJ). **(B)** Number of participants who received at least one trial of LIFU stimulation targeted to the indicated parcel. **(C)** Average of participant-level sham-adjusted non-neutral response rate within the indicated parcel. **(D)** Parcels in which the mean from **(C)** was significant (p<0.05, one-sided t-test, false discovery ratecorrected). **(E)** Cohen’s *d* effect size for each parcel.

### Association of Immediate Cognitive-Emotional Response with Clinical Improvement

In our previous studies we demonstrated significant clinical effects associated with active LIFU sonication [28,29]. Across chronic pain participants included in the present analyses, compared to sham, active LIFU targeted throughout the ACC significantly improved pain measured with the brief pain inventory NRS at 24 hours **(*t* = -3.41, *p* = 0.004)** and seven days post-stimulation **(*t* = -5.05, *p* < 0.001)** (**Fig. 5A**). Pain scores on the PROMIS pain intensity scale were also significantly improved at seven days [28]. Similarly, in the TRD cohort, active LIFU targeted to the sgACC significantly improved mood measured with HDRS-6 at 24 hours following stimulation **(*t* = -2.4, *p* = 0.034)**, but not at seven days **(*t* = -1.23, *p* = 0.24)** (**Fig. 5D**). PANAS-X Sadness subscale scores also significantly improved across the visit [29].

**Figure 5.**
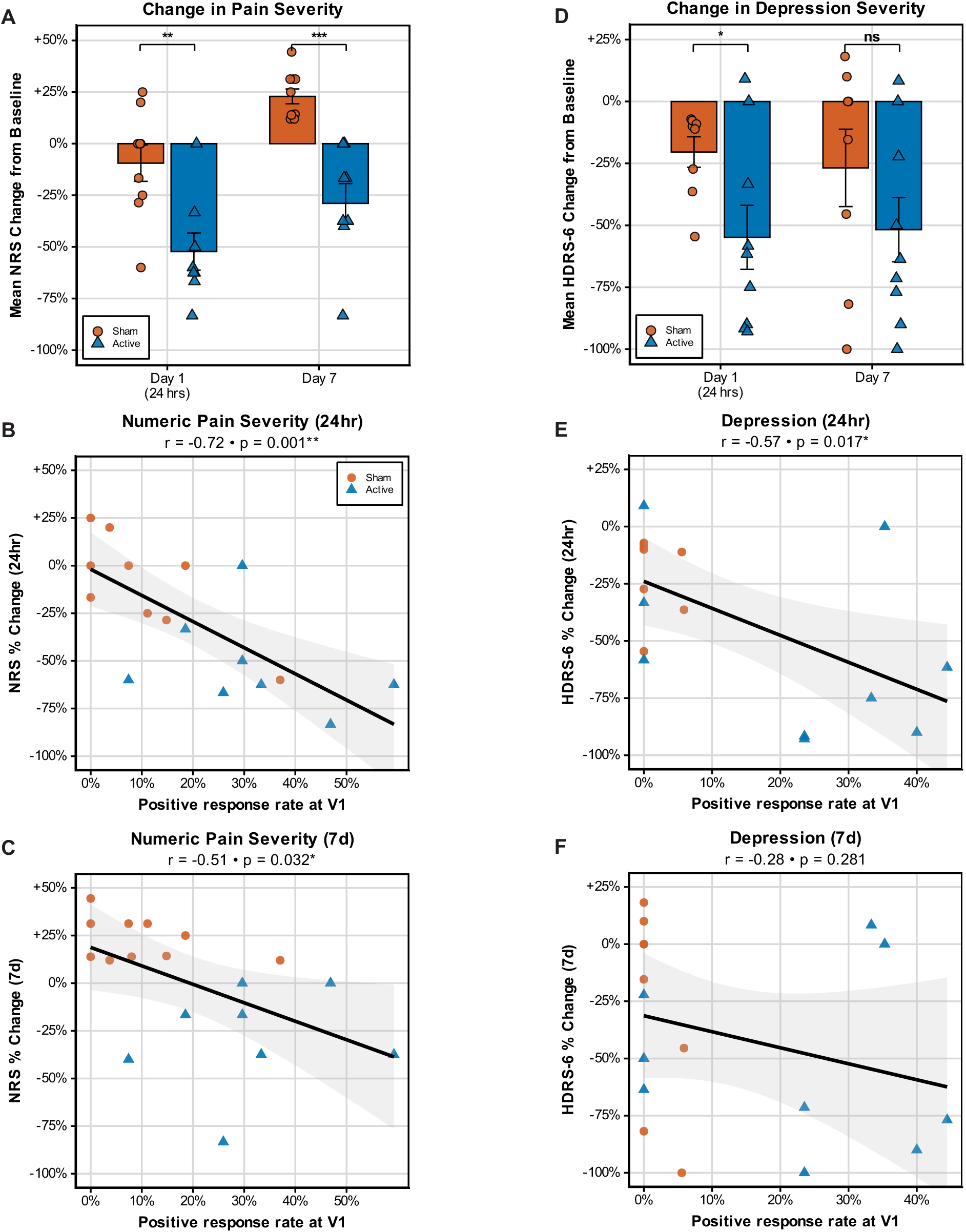
Immediate subjective reports predict subsequent clinical response. Data from **(A-C)** chronic pain and **(D-F)** treatment-resistant depression (TRD) cohorts. **(A)** Change in numeric rating scale (NRS) score from baseline at 24 hours and seven days post-stimulation at visit one (V1) in sham (orange) and active (blue) conditions. Clinical scores along the y-axis are reported as percentage change relative to baseline. Statistics reflect Welch two-sample t-tests comparing sham and active at each timepoint (**\****p*<0.05**, \*\****p*<0.01**, \*\*\****p*<0.001). **(B-C, E-F)** Linear models depicting correlations between positive-valenced immediate subjective response rate at V1 and clinical score changes from **(A)** and **(D)** (same y-axis). Subjective effects on the x-axis reported as percentage of total trials scored as ‘positive’ during the V1 stimulation session. Solid line indicates the least-squares linear regression fit. Pearson correlation coefficients (*r*) and associated two-tailed *p*-values are shown. Shaded regions represent 95% confidence band. **Linear models** for positive response rate versus **(B)** NRS at 24 hours; **(C)** NRS at seven days; **(E)** HDRS-6 at 24 hours; and, **(F)** HRDS-6 at seven days post-stimulation.

Given our findings that active LIFU can produce significant immediate cognitive-emotional effects reportable by individual participants, we sought to determine whether these perceptible immediate responses predict clinically meaningful improvement in pain or depression severity. We hypothesized that the rate of positive immediate response would correlate with subsequent improvement seen on the clinical scales in **Figure 5A, D**.

For the chronic pain cohort, immediate positive-valence response rate at the first stimulation visit correlated significantly with pain severity improvement measured with NRS at both 24 hours (**Fig. 5B**, *r* = −0.72, *p* = 0.001) and seven days (**Fig. 5C**, *r* = −0.51, *p* = 0.032), and marginally with pain intensity measured with PROMIS at seven days (**Fig. S1A**, *r* = −0.47, *p* = 0.051). For the TRD cohort, immediate subjective positive response rate correlated significantly with improvement in HDRS-6 score at 24 hours (**Fig. 5E**, *r* = −0.57, *p* = 0.017) and marginally with improvement in PANAS sadness score during the first stimulation visit (**Fig. S1B**, *r* = −0.46, *p* = 0.071). There was no significant correlation with HDRS-6 score changes at one week follow-up (**Fig. 5F**, *r* = −0.28, *p* = 0.28). These findings provided moderate support for the hypothesis that immediate responses would predict subsequent clinical outcomes.

## DISCUSSION

In the present study, we sought to determine the immediate subjective effects of low-intensity transcranial focused ultrasound (LIFU) delivered to the anterior cingulate cortex (ACC), a key hub for cognitive-emotional integration and regulation of subjective experience [1–3]. Given the ACC’s central role in shaping affective state and its established involvement in chronic pain and treatment-resistant depression (TRD) [1–5], studying ACC-targeted stimulation in these patient populations enabled us to examine the real-time effects of focal neuromodulation within clinically and symptomatically relevant circuitry. Participants underwent repeated brief sham and active LIFU stimulation with successive trial-by-trial reporting of subjective experiential effects, enabling characterization of rapid cognitive-emotional responses to focal neuromodulation at the level of individual stimulation events. We examined how these acute response profiles differed in sham versus active stimulation, how they varied across diagnostic populations and ACC subregion targets, and how they were related to subsequent clinical outcomes. Our findings provide evidence that noninvasive focal modulation of ACC circuitry can produce measurable real-time effects on subjective experience and support the potential utility of immediate patient-reported responses as markers of clinically meaningful neural target engagement.

A primary finding of this study is that active ACC-targeted LIFU elicited immediate subjective effects beyond those observed during sham stimulation. Across both cohorts, active stimulation produced a robust increase in non-neutral responses, particularly positive-valence responses, at both the group level and in a majority of individuals providing direct evidence that focal noninvasive modulation of ACC circuitry can alter subjective experience in real time. Importantly, these effects were observed despite rigorous sham control procedures and auditory masking, supporting the interpretation that the observed responses were not solely attributable to expectancy or nonspecific sensory cues. These findings extend prior work demonstrating aggregate clinical effects of ACC-targeted LIFU [28,29,36] by showing that individual sonications themselves can produce rapid experiential changes perceptible to participants.

At the same time, the predominance of neutral responses and variability across trials highlights the probabilistic and heterogeneous nature of acute neuromodulatory effects. Subjective responses were not uniformly elicited during every stimulation event, and responsiveness varied substantially across individuals. This variability may reflect differences in anatomical targeting precision, baseline circuit state, disease-related neurobiology, or individual sensitivity to neuromodulation [54]. Better understanding of how these factors contribute to the variability we see in elicited subjective experience – and ultimately influence clinical response – will require additional studies focused on characterizing the biological and psychological mediators of ultrasonic neuromodulation. Importantly, the use of open-ended free-response reporting allowed participants to report whichever aspects of stimulation were most salient to them, rather than constraining responses to predefined symptom domains. This approach likely improved sensitivity to subtle or unexpected experiential effects, though it also emphasizes the complexity and multidimensional nature of subjective responses to focal ACC modulation. Effective implementation of cognitive-emotional reports as biomarkers for neuromodulation in clinical settings will require additional studies to define sensitive and specific inquiries which more systematically capture clinically meaningful subjective changes.

The inclusion of two distinct diagnostic populations further provides insight into both shared and disorder-specific aspects of ACC neuromodulation. Despite differences in symptom domain and targeting strategy – distributed ACC sampling in chronic pain versus focused sgACC stimulation in TRD – both cohorts demonstrated increased responsiveness to active stimulation relative to sham. These findings support the concept that LIFU can modulate affective circuitry centered on the ACC across multiple neuropsychiatric conditions. Interestingly, however, the magnitude and distribution of responses differed between cohorts, with the chronic pain cohort showing significantly higher non-neutral sham response rates and the TRD cohort exhibiting a larger active-versus-sham separation in non-neutral response rates. While different neuropsychiatric conditions are associated with differing magnitude of placebo effect [55], more work is needed to systematically characterize these differences and determine whether our observations generalize to other neuromodulation paradigms within these populations. Nonetheless, these differences may reflect distinct underlying pathophysiology, varying baseline affective states, or differences in stimulation focality and sampling density across ACC subdivisions.

Beyond demonstrating immediate subjective effects, another key contribution of this work is the observed relationship between acute experiential responses and subsequent clinical outcomes. Across both cohorts, individuals with higher rates of positive trial-level responses during stimulation demonstrated greater improvement on validated clinical outcome measures, particularly at early follow-up time points. In chronic pain participants, positive response rates strongly predicted reductions in pain severity at both 24 hours and seven days, while in TRD participants, positive responses were associated with improvement in depressive symptoms at 24 hours and showed a trend-level relationship with acute change in Sadness immediately following the stimulation session. These findings suggest that acute positive responses may index effective engagement of symptom-relevant circuitry.

The relationship between acute subjective effects and downstream therapeutic response parallels observations from invasive neuromodulation paradigms such as deep brain stimulation (DBS), where intraoperative patient-reported effects can be used to refine targeting and programming and, in some cases, may predict long-term clinical effects [37–39]. For example, patient-reported feedback in DBS treatment of essential tremor enables effective modulation of causal disease circuitry with lower amplitudes and subjectively favorable response profiles [37]. Improved subjective outcomes are similarly seen in DBS treatment of Parkinson’s disease [38]. These studies emphasize patient-reported measures for minimizing negative side effects and focus on predominantly physical symptoms. In DBS for depression, 3-minute stimulation trials targeting variable sgACC subregions elicited variable immediate subjective responses, with the ‘best’ subjective response in all patients sharing a common anatomical stimulation target known to be optimal for clinical effect [39,56]. Collectively, these DBS results demonstrate clinically relevant efficacy of patient-reported feedback in guiding neuromodulation despite their noted trial-level variability at the subject-level. The present findings extend this principle to a similarly focal yet noninvasive modality, raising the possibility that real-time subjective feedback during LIFU could be used to select optimal ultrasonic stimulation parameters. Such feedback could also inform individualized targeting for subsequent treatment sessions. Systematic interrogation of ACC subregions might even be developed into a standardized diagnostic assessment to identify circuit-defined biotypes for depression or chronic pain. Such an approach may be particularly important given the known functional heterogeneity of ACC subregions and the network-level heterogeneity observed across affective disorders [57,58].

Several limitations should be considered. First, subjective responses were collected by an unblinded device operator. Although this approach was deliberate to optimize participant safety (i.e. avoiding targets associated with negative responses), we cannot rule out the possibility that recording of participant responses was biased by lack of operator blinding. Such bias could inflate the odds ratios measured for active versus sham comparisons. Several observations argue against pervasive bias: many individuals showed an odds ratio near or even less than 1; substantially different responses were found for the two concurrent studies (chronic pain and TRD); and responsiveness varied across ACC subregions. A second limitation is related to the collection of open-ended self-reports and subsequent categorization by a blinded rater. While this approach captured rich experiential data, it introduced potential variability in interpretation and lacked the external validity and granularity of standardized quantitative rating scales. Future studies should incorporate structured, blinded survey methods capable of capturing rapid subjective effects prospectively and with greater dimensional specificity. Third, although sham stimulation included auditory masking, and prior work demonstrated preserved blinding integrity in the TRD cohort [29], subtle perceptual differences between active and sham conditions cannot be completely excluded [59]. Fourth, analyses relating acute responses to clinical outcomes were restricted to the initial treatment visit. This was done deliberately to minimize functional unblinding and to account for many participants who had significant improvement on clinical scales rendering their baseline second-visit scores too low to reliably assess further improvement. While this approach may limit generalizability across repeated treatment sessions, it underscores the potential impact of employing a feedback-driven approach early in neuromodulation regimes to optimize subsequent clinical response. Finally, sample sizes were modest for some subgroup and subject-level analyses, limiting the power to detect true effects and potentially inflating effect sizes. Larger studies will be needed to further characterize sources of inter-individual variability and refine predictive biomarkers of response.

Despite these limitations, the present study provides evidence that brief ACC-targeted LIFU stimulation can induce rapid, trial-specific changes in subjective cognitive-emotional state that distinguish active from sham stimulation and relate to subsequent clinical improvement. These findings help bridge a critical gap between noninvasive neural target engagement and patient-reported experience, supporting the potential utility of immediate subjective responses as biomarkers of effective neuromodulation. More broadly, this work highlights the promise of LIFU as a tool for causally probing ACC function and for developing personalized, circuit-guided neuromodulation strategies for neuropsychiatric disorders.

## Data Availability

All data produced in the present study are available upon reasonable request to the authors.

