## Supplemental Figure 1 for "Cognitive-emotional responses to ultrasonic neuromodulation of anterior cingulate cortex"

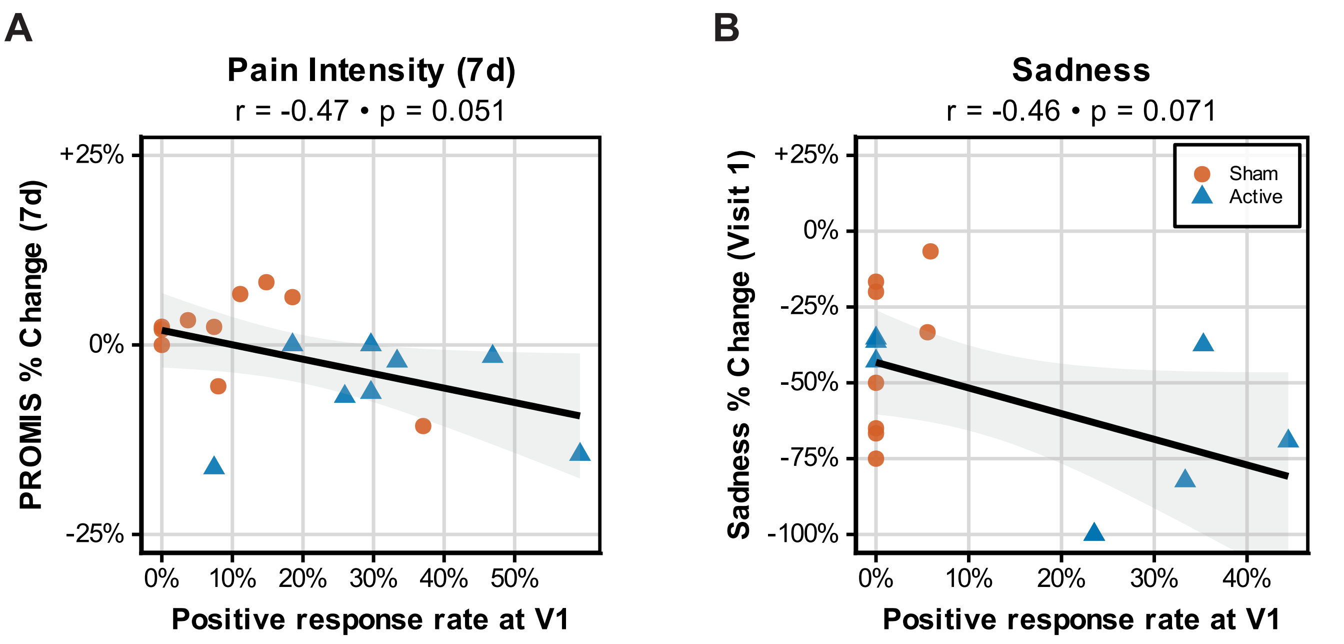


**Supplementary Figure 1. Immediate subjective reports marginally predict subsequent pain intensity and sadness improvement**. Linear models depicting correlations between positive-valenced immediate subjective response rate at visit one (V1) and clinical score changes in sham (orange) and active (blue) conditions. Immediate subjective responses versus **(A)** Patient-Reported Outcomes Measurement Information System (PROMIS) pain intensity score changes at seven days post-stimulation among chronic pain participants; and, **(B)** Positive and Negative Affect Schedule Expanded (PANAS-x) sadness subscale scores from pre to post-stimulation among treatment-resistant depression (TRD) participants. Subjective effects on the x-axis reported as percentage of total trials scored as 'positive' during the V1 stimulation session. Clinical scores along the y-axis are reported as percentage change relative to baseline. Solid line indicates the least-squares linear regression fit. Pearson correlation coefficients (*r*) and associated two-tailed *p*-values are shown. Shaded regions represent 95% confidence band.
